# Prospective predictive performance comparison between Clinical Gestalt and validated COVID-19 mortality scores

**DOI:** 10.1101/2021.04.16.21255647

**Authors:** Adrian Soto-Mota, Braulio A. Marfil-Garza, Santiago Castiello de Obeso, Erick Martínez, Daniel Alberto Carrillo-Vázquez, Hiram Tadeo-Espinoza, Jessica Paola Guerrero-Cabrera, Francisco Eduardo Dardón-Fierro, Juan Manuel Escobar Valderrama, Jorge Alanis-Mendizabal, Juan Gutiérrez

**Affiliations:** National Institute of Medical Sciences and Nutrition Salvador Zubirán; The University of Oxford

**Author notes:** **Corresponding author:** Luis Adrian Soto-Mota MD, DPhil. Assistant Professor at the Metabolic Diseases Research Unit of The National Institute of Medical Sciences and Nutrition Salvador Zubirán. Mexico City, Postal code: 14080.

**Keywords:** COVID-19, Clinical Gestalt, NEWS2, LOW-HARM, qSOFA, Nutri-Cov

## Abstract

**Background:** Most COVID-19 mortality scores were developed in the early months of the pandemic and now available evidence-based interventions have helped reduce its lethality. It has not been evaluated if the original predictive performance of these scores holds true nor compared it against Clinical Gestalt predictions. We tested the current predictive accuracy of six COVID-19 scores and compared it with Clinical Gestalt predictions.

**Methods:** 200 COVID-19 patients were enrolled in a tertiary hospital in Mexico City between September and December 2020. Clinical Gestalt predictions of death (as a percentage) and LOW-HARM, qSOFA, MSL-COVID-19, NUTRI-CoV and NEWS2 were obtained at admission. We calculated the AUC of each score and compared it against Clinical Gestalt predictions and against their respective originally reported value.

**Results:** 106 men and 60 women aged 56+/-9 and with confirmed COVID-19 were included in the analysis. The observed AUC of all scores was significantly lower than originally reported; LOW-HARM 0.96 (0.94-0.98) vs 0.76 (0.69-0.84), qSOFA 0.74 (0.65-0.81) vs 0.61 (0.53-0.69), MSL-COVID-19 0.72 (0.69-0.75) vs 0.64 (0.55-0.73) NUTRI-CoV 0.79 (0.76-0.82) vs 0.60 (0.51-0.69), NEWS2 0.84 (0.79-0.90) vs 0.65 (0.56-0.75), Neutrophil-Lymphocyte ratio 0.74 (0.62-0.85) vs 0.65 (0.57-0.73). Clinical Gestalt predictions were non-inferior to mortality scores (AUC=0.68 (0.59-0.77)). Adjusting the LOW-HARM score with locally derived likelihood ratios did not improve its performance. However, some scores performed better than Clinical Gestalt predictions when clinician’s confidence of prediction was <80%.

**Conclusion:** No score was significantly better than Clinical Gestalt predictions. Despite its subjective nature, Clinical Gestalt has relevant advantages for predicting COVID-19 clinical outcomes.

## INTRODUCTION

Many prediction models have been developed for COVID-19 (1–5) and their applications in healthcare range from bed-side counseling to triage systems (6). However, most have been developed within specific clinical contexts (1,2) or validated with data from the early months of the pandemic (4,5). Since then, health systems have implemented protocols and adaptations to cope with a surge in hospitalization rates (7), and now, clinicians have more knowledge and experience for managing these patients. Additionally, other non-biological factors like critical-care availability have been found to strongly influence the prognosis of COVID-19 patients (8,9). These frequently intangible factors (e.g., the experience of the staff with specific healthcare tasks) impact prognosis but are ignored by mortality scores.

Prediction models are context-sensitive (10), therefore, to preserve their accuracy they must be applied in contexts as similar as possible to the ones where they were derived from. Considering that healthcare systems and settings are quite different around the world, there are many examples of scores requiring adjustments or local adaptations (11,12).

Predicting is an every-day activity in most medical fields and, in other scenarios, clinician’s subjective predictions have been observed to be as accurate as mathematically derived models (13–15). However, the opposite has been observed as well, for example, clinicians tend to overestimate the long-term survival of oncologic patients (16). This work aimed to compare the predictive performance of different mortality prediction models for COVID-19 (some of them in the same hospital they were developed) against their original performance and Clinical Gestalt predictions.

## METHODS

### Study design

To test the hypothesis that the predictive performance of already validated scores declined over time, we chose the LOW-HARM (4), MSL-COVID-19, and NUTRI-CoV (5) scores because all three were validated with data from Mexican COVID-19 patients. To rule out that this was a phenomenon exclusive of scores developed with Mexican data, we reevaluated the accuracy of the NEWS2 (1) and qSOFA (2) scores, and the Neutrophil:Lymphocyte ratio to predict mortality from COVID-19(17).

Clinical Gestalt predictions and all necessary data to calculate the prognostic scores were obtained at hospital admission from October to December 2020. The Internal Medicine residents in charge of collecting the clinical history, physical examination and the initial imaging and laboratory workup were asked after their all the initial imaging and laboratory reports were available:

1. How likely do you think it is this patient will die from COVID-19? (as a percentage).
2. How confident are you of that prediction? (as a percentage).

Additionally, to test the hypothesis that updating the statistical weights of a score with local data could help preserve its original accuracy, we developed a second version of the LOW-HARM score (LOW-HARM score v2) using positive and negative likelihood ratios derived from cohorts of Mexican patients (4,8) (instead of only positive likelihood ratios from Chinese patients (18,19) as in the original version).

The likelihood ratios (LR+/LR-) used to calculate the LOW-HARM score v2 were as follows: oxygen saturation <88% = 2.61/0.07, previous diagnosis of hypertension = 2.37/0.65, elevated troponin (>20 pg/mL) =15.6/0.62, elevated CPK (>223 U/L) =2.37/0.88, leukocyte count > 10 000 cells/μL = 5.6/0.48, lymphocyte count <800 cells/μL (<0.8 cells/mm3) = 2.24/0.48, serum creatinine > 1.5 mg/dL = 19.1/0.6.

Finally, to test the hypothesis that scores outperformed Clinical Gestalt predictions when their confidence was “low” (below or equal to the median perceived confidence (i.e., < 80%), we conducted a comparative AUC analysis of cases below or above this threshold.

This study was approved by the Ethics Committee for Research on Humans of the National Institute of Medical Sciences and Nutrition Salvador Zubirán on August 25, 2020 (Reg. No. DMC-3369-20-20-1-1a).

### Sample size rationale

We calculated with “easyROC” (20), an open R-based web-tool for estimating sample sizes for AUC direct and non-inferior comparisons using Obuchowski’s method (21) that; for detecting no-inferiority with a >0.05 maximal AUC difference with the reported LOW-HARM AUC (0.96 95% CI:0.94 – 0.98) with a case allocation ratio of 0.7 (because the mortality in our centre is ∼ 0.3) with a power of 0.8 and a significance cut-off level of 0.05, 159 patients would be needed. For detecting >0.1 differences between AUCs, 99 patients are needed with the rest of the parameters held constant. To allow a patient-loss rate of ∼ 25%, we obtained data from 200 consecutive hospital admissions.

### Setting

A tertiary hospital in Mexico City, fully dedicated to COVID-19 healthcare between October and December 2020.

### Selection of participants

Data from 200 consecutive hospital admissions (with an RT-PCR confirmed COVID-19 infection) were obtained between October and December of 2020. We excluded from the analysis all patients without a documented clinical outcome (e.g., hospitalized at the moment of data collection, transferred to another hospital, voluntary discharge). A total of 166 patients were included in the analysis because 34 patients were either transferred to other hospitals or voluntarily discharged.

### Analysis

Clinical and demographic data were analysed using mean or median (depending on their distribution) and standard deviation or interquartile range (IQR) as dispersion measures. Shapiro-Wilk tests were used for assessing if variables were normally distributed.

R version 4.0.3 using the packages “caret” for confusion matrix calculations, “pROC” for ROC analysis, and STATA v12 software were used for statistical analysis. The AUCs differences were analysed using DeLong’s method with the STATA function “roccomp” (22). A p value of <0.05 for inferring statistical significance was for all statistical tests.

## RESULTS

We include 166 patients in our study. Of these, 47 (28.3%) were deaths and 119 (71.7%) were survivors. General demographics and clinical characteristics of these populations are shown in Table 1. As expected, decreased peripheral saturation, ventilatory support, cardiac injury, renal injury, leukocytosis, and lymphocytosis were more prevalent in the group of patients that died during their hospitalization.

**Table 1.** Patient demographics and clinical data.

|  | **Total (n=166)** | **Died (n=47)** | **Survived (n=119)** | **P\*** |
| --- | --- | --- | --- | --- |
| **Female, n (%)** | 60 (36.1) | 18 (38.3) | 42 (35.3) | 0.717 |
| **Age, years (IQR)** | 56 (45-64) | 61 (54-69) | 52 (42-63) | 0.0002 |
| **Weight, kg (IQR)** | 78 (70-90) | 78 (65-96) | 79 (72-90) | 0.659 |
| **Height, cm (IQR)** | 165 (158-170) | 165 (160-172) | 164 (580-170) | 0.578 |
| **BMI (IQR)** | 29 (25.4-33) | 28 (24-33) | 29 (27-32) | 0.302 |
| **Obesity, n (%)** | 77 (46.4) | 21 (44.7) | 56 (47.1) | 0.782 |
| **Diabetes Mellitus, n (%)** | 42 (25.3) | 12 (25.5) | 30 (25.2) | 0.966 |
| **Hypertension, n (%)** | 49 (29.5) | 17 (36.2) | 32 (26.9) | 0.238 |
| **Smoking, n (%)** | 37 (22.3) | 10 (21.3) | 27 (22.7) | 0.844 |
| **Immunosuppression, n (%)** | 25 (15.1) | 6 (12.8) | 19 (16.0) | 0.603 |
| **COPD, n (%)** | 7 (4.2) | 4 (8.5) | 3 (2.5) | 0.084 |
| **CKD, n (%)** | 9 (5.4) | 2 (4.3) | 7 (5.9) | 0.677 |
| **CAD, n (%)** | 8 (4.8) | 4 (8.5) | 4 (3.4) | 0.163 |
| **SpO2% <88%, n (%)** | 156 (94.0) | 47 (100) | 109 (91.6) | 0.040 |
| **IMV/CPAP, n (%)** | 96 (57.8) | 33 (70.2) | 63 (52.9) | 0.042 |
| **Positive troponin/CPK, n (%)** | 77 (46.4) | 31 (66) | 46 (38.7) | 0.001 |
| **Creatinine >1.5 mg/dL, n (%)** | 25 (15.1) | 14 (29.8) | 11 (9.2) | 0.001 |
| **WBC >10,000 cells/ μL, n (%)** | 94 (56.6) | 35 (74.5) | 59 (50) | 0.004 |
| **Lymphocytes <800 cells/μL, n (%)** | 113 (68.1) | 38 (80.9) | 75 (63) | 0.026 |
| **Neutrophil:lymphocyte ratio >9.8 (%)** | 60 (36.1) | 27 (57.5) | 33 (27.7) | <0.0001 |
| **Length-of-stay, days (IQR)** | 15.5 (9-27) | 17 (11-27) | 13 (8-27) | 0.5408 |

BMI: body-mass index, COPD: chronic obstructive pulmonary disease, CKD: chronic kidney disease, CAD: coronary artery disease,
CPK: creatine phosphokinase, WBC: white blood cells, IMV: invasive mechanical ventilation, CPAP: continuous positive airway pressure.
\* Comparisons were done between deaths and survivors. $\chi^2$ was used to compare categorical variables, Mann-Whitney U test was used to compare continuous variables.

Table 2 shows the median scores and their IQR for each prediction tool. As expected, there was a more pronounced mean difference between groups in scores that were based on a 100-point scale (clinical gestalt, LOW-HARM scores). Table 2 shows the originally reported AUC vs the AUC we observed in our data.

**Table 2.** Distribution and accuracy of selected mortality prediction tools.

| Prediction tool | Total<br>(n=166) | Died<br>(n=47) | Survived<br>(n=119) | Original AUC (95% CI) | Observed AUC (95% CI) |
| --- | --- | --- | --- | --- | --- |
| Clinical Gestalt –<br>Confidence (IQR) | 30 (20-50)<br>80 (70-90) | 40 (30-70)<br>80 (60-90) | 30 (15-40)<br>80 (70-90) | - | 0.68 (0.59 - 0.77) |
| LOW-HARM (IQR) | 46 (8.4-83.8) | 86 (37.5-99.3) | 37.5 (6.4-69) | 0.96 (0.94 - 0.98) | 0.76 (0.69 - 0.84) |
| LOW-HARM v2 (IQR) | 9.7 (0.9-52.7) | 49 (9.7-96.3) | 3.2 (0.5-28.1) | - | 0.78 (0.71 - 0.86) |
| NUTRI-CoV (IQR) | 9 (7-12) | 10 (8-12) | 9 (7-11) | 0.79 (0.76 - 0.82) | 0.60 (0.51 - 0.69) |
| MSL-COVID-19 (IQR) | 8 (7-10) | 8 (8-10) | 8 (7-9) | 0.72 (0.69 - 0.75) | 0.64 (0.55 - 0.73) |
| qSOFA (IQR) | 1 (1-1) | 1 (1-2) | 1 (1-1) | 0.74 (0.65 - 0.81) | 0.61 (0.53 - 0.69) |
| NEWS2 (IQR) | 7.5 (6-9) | 9 (7-10) | 7 (5-9) | 0.84 (0.79 - 0.90) | 0.65 (0.56 - 0.75) |
| Neutrophil:<br>Lymphocyte ratio >9.8<br>(%) | 64.4 | 55.3 | 27.7 | 0.74 (0.62 - 0.85) | 0.65 (0.57 - 0.73) |

\*To calculate the relative mean difference, some scores (those not based in 100 points), were converted to a percentage in the following manner: (patient score/maximum possible score) \*100.
Overall comparison test for observed AUC=0.002
Individual AUROC comparisons:
- Clinical Gestalt vs all scores, $p>0.05$

### Performance characteristics of selected predictive models and AUC comparisons

Figure 1 shows the performance characteristics of the selected predictive models. Overall, we found a statistically significant difference between predictive models (p=0.002).

**Figure 1.**
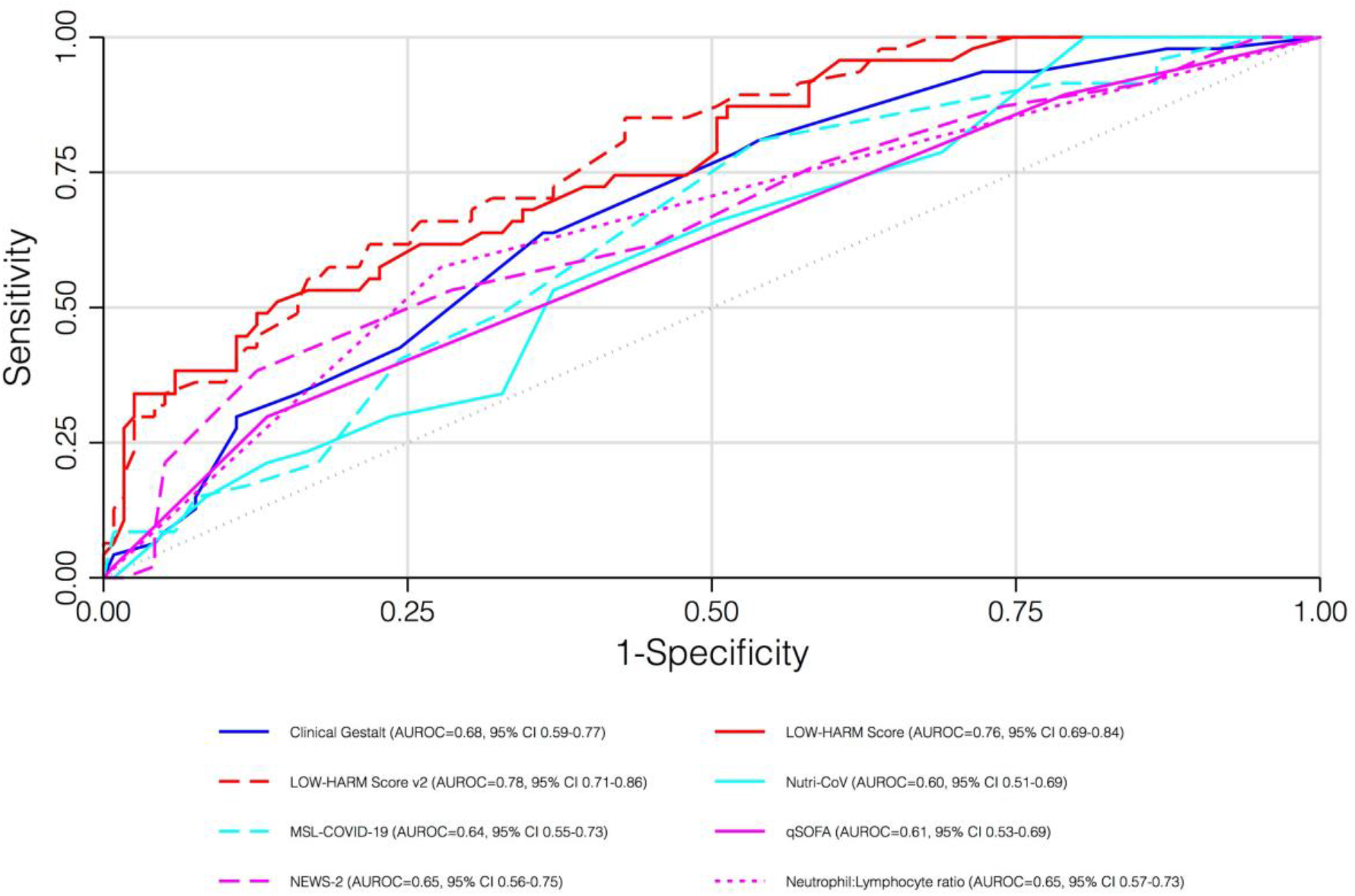
AUC comparison for selected mortality prediction tools.

However, we did not find statistically significant differences between Clinical Gestalt and other prediction tools.

As expected, we found that the confidence of prediction increased in cases in which the predicted probability of death was clearly high or clearly low (Figure 2). We found a moderate-strong, bimodal, correlation between the confidence of prediction and the predicted probability of death at a <50% predicted probability of death (Pearson’s R=0.60, p<0.0001) and at a >50% predicted probability of death (Pearson’s R=0.50, p=0.0002).

**Figure 2.**
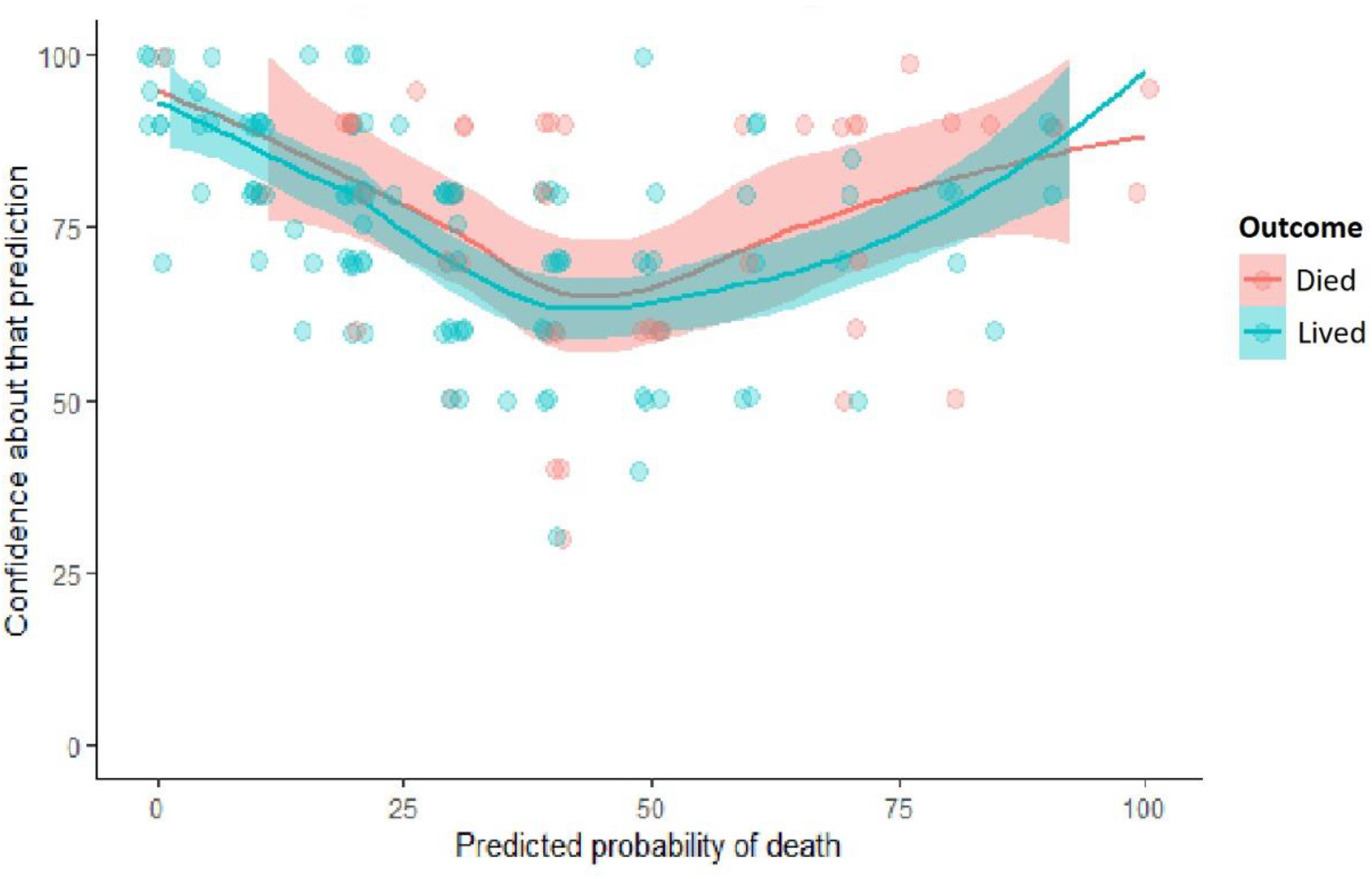
Clinical gestalt prediction and confidence of prediction.

We further explored the performance characteristics of the selected predictive models in specific contexts (Appendix Table 1). Figure 3 shows the results of the analysis including cases in which the certainty of prediction was below and above 80%. Overall, we found a statistically significant difference between predictive models in both settings. In cases in which the confidence of prediction was < 80%, both versions of the LOW-HARM scores showed a larger AUC compared to Clinical Gestalt (Figure 3b and Appendix Table 1). An additional analysis restricted to cases in which the certainty of prediction was <80% and the predicted probability of death was <30% (i.e., median value for all cases) found a statistically significant difference between predictive models (p=0.0005). Similarly, individual comparisons showed a larger AUC statistically significant differences between Clinical Gestalt and both versions of the LOW-HARM score (Appendix Table 1).

**Figure 3.**
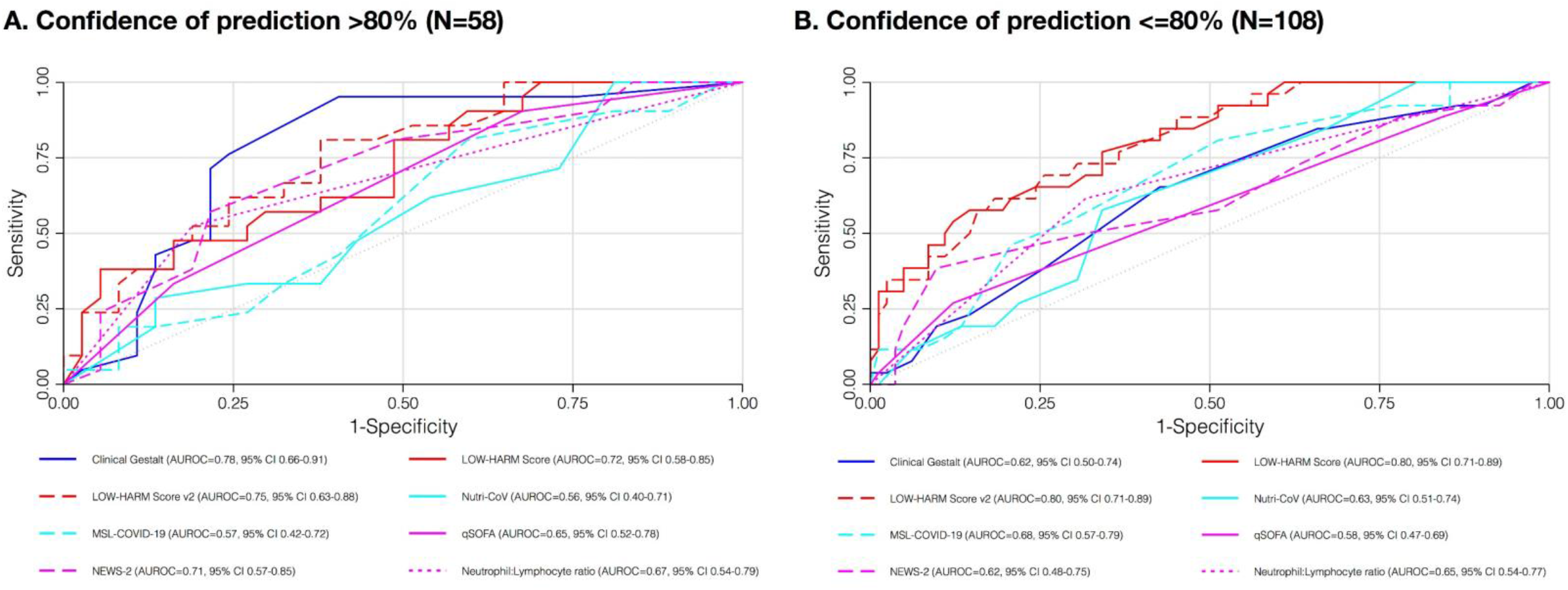
AUC comparison for selected mortality prediction tools according to the confidence of prediction. Panel A shows the AUC comparison for selected mortality prediction tools in cases where the confidence of prediction was >80%. Panel B shows the AUC comparison for selected mortality prediction tools in cases where the confidence of prediction was <80%.

## DISCUSSION

Outcome prediction plays an important role in every-day clinical practice. This work highlights the inherent limitations of statistically derived scores and some of the advantages of Clinical Gestalt predictions. In other scenarios where using predictive scores is frequent, more experienced clinicians can always ponder their sometimes subjective yet, quite valuable insight. However, with the COVID-19 pandemic clinicians with all levels of training started their learning curve at the same time. In this study, we had the unique opportunity of re-evaluating more than one score (two of them in the same setting and for the same purpose they were designed for), while testing the accuracy of Clinical Gestalt, in a group of clinicians who started their learning curve for managing a disease at the same time (experience and training withing healthcare teams is usually mixed for other diseases).

Additionally, we explored the accuracy of Clinical Gestalt across different degrees of prediction confidence. To our knowledge, this is the first time that this type of analysis is done for subjective clinical predictions and proved to be quite insightful. The fact that Clinical Gestalt’s accuracy correlates with confidence in prediction, suggests that while there is value in subjective predictions, it is also important to ask ourselves about how confident we are about our predictions. Interestingly, our results suggest Clinical Gestalt predictions are particularly prone to be positively biased, clinicians were more likely to correctly predict which patients would survive than which patients would die (Figure 2 and Supplementary Figure 1). This is consistent with other studies that have found that clinicians tend to overestimate the effectiveness of their treatments and therefore, patient survival (16).

Since it is expected that scores will lose at least some of their predictive accuracy when used outside the context they were developed in, it has already been reported that local adaptations improve or help retain their predictive performance. In this work, we tried to evaluate if by updating the likelihood ratio values used in the calculation of the LOW-HARM score with data from Mexican patients we could mitigate its loss of accuracy. However, despite the AUC of the LOW-HARM score v2 being slightly larger than the AUC of the original LOW-HARM score, the difference was not statistically significant nor significantly more accurate than Clinical Gestalt predictions. This highlights the fact that scores are far from being final or perfect tools even after implementing local adjustments.

Even when some of the results in this study can prove insightful for other clinical settings and challenges, our results cannot be widely extrapolated due to the local setting of our work and the highly heterogenous nature of COVID-19 healthcare systems. Additionally, it is likely that emerging variants, vaccination, or the seasonality of the contagion waves (23) will continue to influence the predictive capabilities of all predictive models. Additionally, our sample size was calculated to detect non-inferiority between prediction methods. Specifically designed studies are needed to better investigate the relationship between subjective confidence, accuracy, and positive bias.

Clinical predictions will always be challenging because all medical fields are in constant development and clinical challenges are highly dynamic phenomena. Despite its inherent subjectivity, Clinical Gestalt immediately incorporates context specific factors and, in contrast with statistically derived models, it is likely to improve its accuracy over time.

## CONCLUSION

All scores had lower predictive accuracy than in their original publications. No score showed better predictive performance than Clinical Gestalt predictions however, scores could still outperform Clinical Gestalt when confidence in Clinical Gestalt predictions is perceived to be low. Prognostic scores require constant reevaluation even after being properly validated and adjusted. No score can or should ever substitute careful medical assessments and thoughtful clinical judgement.

## Data Availability

Anonymized data will be shared upon request to the corresponding author.

## AUTHORS CONTRIBUTION

All authors contributed significantly to the design analysis and reporting of this study. Dr Adrian Soto-Mota is the guarantor of this study and takes responsibility for the contents of this article.

## ACKNOWLEDGEMENTS

Dr. Braulio A. Marfil-Garza is currently supported by the Patronage of the National Institute of Medical Sciences and Nutrition Salvador Zubirán by the Foundation for Health and Education Dr. Salvador Zubirán (FunSaEd), and the CHRISTUS Excellence and Innovation Center. All authors wish to thank the invaluable support of the National Institute of Medical Sciences and Nutrition Salvador Zubirán Emergency Department staff.

---

**Supplementary figure 1.**
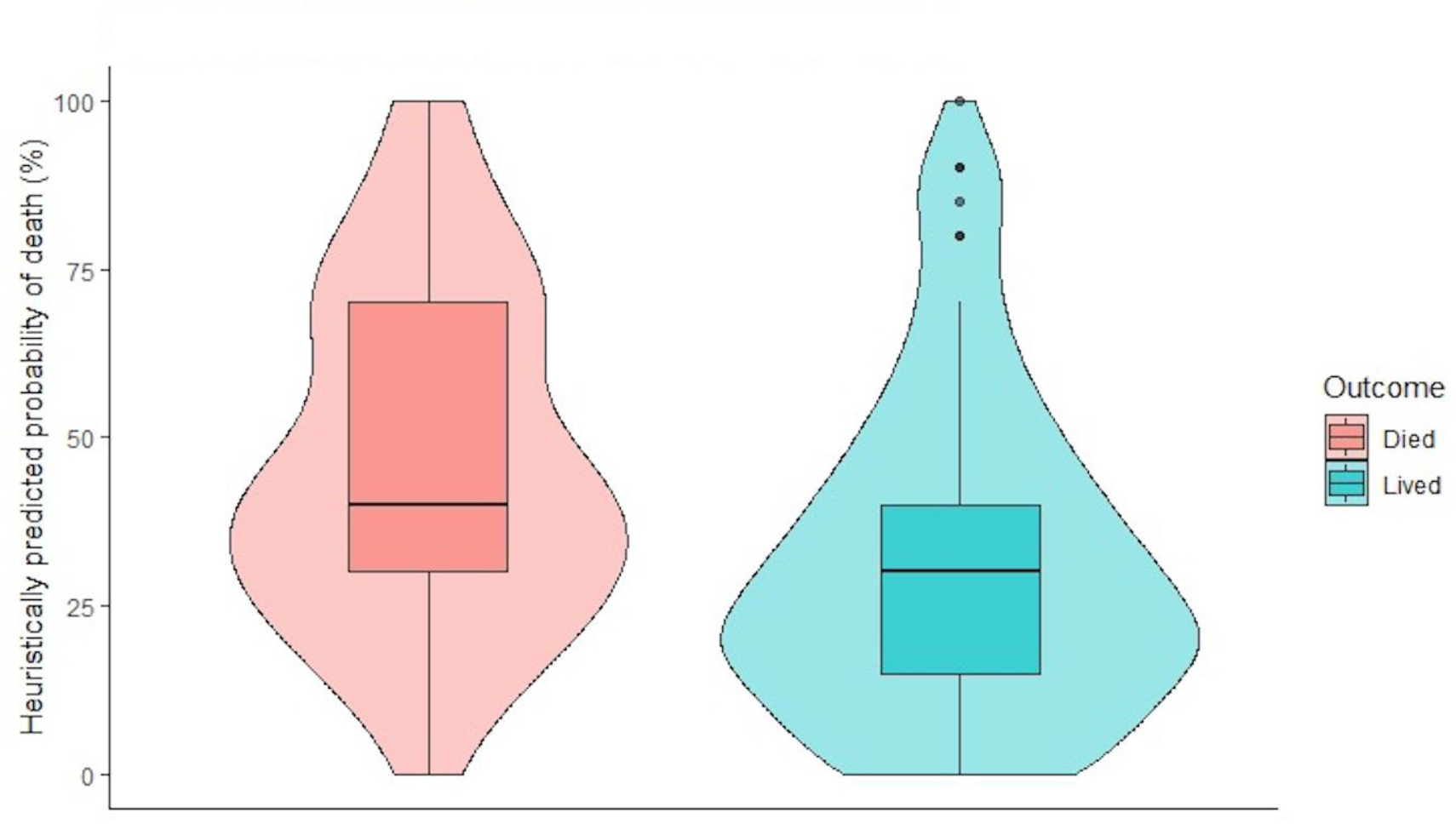
Heuristically predicted probability of death.

## REFERENCES

1. Rigoni M, Torri E, Nollo G, Delle Donne L, Cozzio S. NEWS2 is a valuable tool for appropriate clinical management of COVID-19 patients [Internet]. Vol. 85, European Journal of Internal Medicine. Elsevier B.V.; 2021 [cited 2021 Mar 31]. p. 118–20. Available from: https://doi.org/10.1016/j.ejim.2020.11.020

2. Liu S, Yao N, Qiu Y, He C. Predictive performance of SOFA and qSOFA for in-hospital mortality in severe novel coronavirus disease. American Journal of Emergency Medicine. 2020 Oct 1;38(10):2074–80.

3. Ma A, Cheng J, Yang J, Dong M, Liao X, Kang Y. Neutrophil-to-lymphocyte ratio as a predictive biomarker for moderate-severe ARDS in severe COVID-19 patients [Internet]. Vol. 24, Critical Care. BioMed Central; 2020 [cited 2021 Mar 22]. p. 288. Available from: https://ccforum.biomedcentral.com/articles/10.1186/s13054-020-03007-0

4. Soto-Mota A, Marfil-Garza BA, Martínez Rodríguez E, Barreto Rodríguez JO, López Romo AE, Alberti Minutti P, et al. The low-harm score for predicting mortality in patients diagnosed with COVID-19: A multicentric validation study. Journal of the American College of Emergency Physicians Open [Internet]. 2020 Oct 15;1(6):1436–43. Available from: https://onlinelibrary.wiley.com/doi/10.1002/emp2.12259

5. Bello-Chavolla OY, Antonio-Villa NE, Ortiz-Brizuela E, Vargas-Vázquez A, Gonzále z-Lara MF, de Leon AP, et al. Validation and repurposing of the MSL-COVID-19 score for prediction of severe COVID-19 using simple clinical predictors in a triage setting: The Nutri-CoV score. Ashkenazi I, editor. PLOS ONE [Internet]. 2020 Dec 16 [cited 2021 Apr 1];15(12):e0244051. Available from: https://dx.plos.org/10.1371/journal.pone.0244051

6. White DB, Lo B. A Framework for Rationing Ventilators and Critical Care Beds During the COVID-19 Pandemic. JAMA [Internet]. 2020 Mar 27 [cited 2020 Apr 5]; Available from: http://www.ncbi.nlm.nih.gov/pubmed/32219367

7. OECD/European Union. How resilient have European health systems been to the COVID-19 crisis? In: Health at a Glance: Europe 2020: State of Health in the EU Cycle. 2020. p. 23–81.

8. Olivas-Martínez A, Cárdenas-Fragoso JL, Jiménez JV, Lozano-Cruz OA, Ortiz-Brizuela E, Tovar-Méndez VH, et al. In-hospital mortality from severe COVID-19 in a tertiary care center in Mexico City; causes of death, risk factors and the impact of hospital saturation. Lazzeri C, editor. PLOS ONE [Internet]. 2021 Feb 3 [cited 2021 Mar 22];16(2):e0245772. Available from: https://dx.plos.org/10.1371/journal.pone.0245772

9. Najera H, Ortega-Avila AG. Health and Institutional Risk Factors of COVID-19 Mortality in Mexico, 2020. American Journal of Preventive Medicine [Internet]. 2021 Apr 1 [cited 2021 Apr 4];60(4):471–7. Available from: https://doi.org/10.1016/j.amepre.2020.10.015

10. Khan Z, Hulme J, Sherwood N. An assessment of the validity of SOFA score based triage in H1N1 critically ill patients during an influenza pandemic. Anaesthesia [Internet]. 2009 Dec 1 [cited 2020 May 24];64(12):1283–8. Available from: http://doi.wiley.com/10.1111/j.1365-2044.2009.06135.x

11. Fronczek J, Polok K, Devereaux PJ, Górka J, Archbold RA, Biccard B, et al. External validation of the Revised Cardiac Risk Index and National Surgical Quality Improvement Program Myocardial Infarction and Cardiac Arrest calculator in noncardiac vascular surgery. British Journal of Anaesthesia [Internet]. 2019 Oct 1 [cited 2021 Apr 1];123(4):421–9. Available from: http://bjanaesthesia.org/article/S0007091219304337/fulltext

12. Carr E, Bendayan R, Bean D, Stammers M, Wang W, Zhang H, et al. Evaluation and improvement of the National Early Warning Score (NEWS2) for COVID-19: a multi-hospital study. BMC Medicine [Internet]. 2021 Dec 1 [cited 2021 Apr 1];19(1):23. Available from: https://bmcmedicine.biomedcentral.com/articles/10.1186/s12916-020-01893-3

13. Ros MM, van der Zaag-Loonen HJ, Hofhuis JGM, Spronk PE. SURvival PRediction In SEverely Ill Patients Study—The Prediction of Survival in Critically Ill Patients by ICU Physicians. Critical Care Explorations [Internet]. 2021 Jan 11 [cited 2021 Apr 1];3(1):e0317. Available from: https://journals.lww.com/10.1097/CCE.0000000000000317

14. Donzé J, Rodondi N, Waeber G, Monney P, Cornuz J, Aujesky D. Scores to predict major bleeding risk during oral anticoagulation therapy: A prospective validation study. American Journal of Medicine. 2012 Nov 1;125(11):1095–102.

15. Nazerian P, Morello F, Prota A, Betti L, Lupia E, Apruzzese L, et al. Diagnostic accuracy of physician’s gestalt in suspected COVID-19: Prospective bicentric study. Academic Emergency Medicine. 2021;(February):1–8.

16. Cheon S, Agarwal A, Popovic M, Milakovic M, Lam M, Fu W, et al. The accuracy of clinicians’ predictions of survival in advanced cancer: A review. Vol. 5, Annals of Palliative Medicine. AME Publishing Company; 2016. p. 22–9.

17. Liu J, Liu Y, Xiang P, Pu L, Xiong H, Li C, et al. Neutrophil-to-Lymphocyte Ratio Predicts Severe Illness Patients with 2019 Novel Coronavirus in the Early Stage. medR [Internet]. 2020 [cited 2020 May 24]; Available from: https://doi.org/10.1101/2020.02.10.20021584

18. Zhou F, Yu T, Du R, Fan G, Liu Y, Liu Z, et al. Clinical course and risk factors for mortality of adult inpatients with COVID-19 in Wuhan, China: a retrospective cohort study. The Lancet. 2020 Mar 28;395(10229):1054–62.

19. Yan L, Zhang H-T, Goncalves J, Xiao Y, Wang M, Guo Y, et al. An interpretable mortality prediction model for COVID-19 patients. Nature Machine Intelligence. 2020 May 14;2(5):283–8.

20. Goksuluk D, Korkmaz S, Zararsiz G, Karaagaoglu AE. EasyROC: An interactive web-tool for roc curve analysis using r language environment. R Journal. 2016;8(2):213–30.

21. Obuchowski NA. ROC Analysis. American Journal of Roentgenology [Internet]. 2005 Feb 23 [cited 2021 Apr 1];184(2):364–72. Available from: http://www.ajronline.org/doi/10.2214/ajr.184.2.01840364

22. DeLong ER, DeLong DM, Clarke-Pearson DL. Comparing the Areas under Two or More Correlated Receiver Operating Characteristic Curves: A Nonparametric Approach. Biometrics [Internet]. 1988 Sep [cited 2020 Aug 26];44(3):837. Available from: https://www.jstor.org/stable/2531595

23. Birkmeyer JD, Barnato A, Birkmeyer N, Bessler R, Skinner J. The impact of the COVID-19 pandemic on hospital admissions in the United States. Health Affairs [Internet]. 2020 Nov 1 [cited 2021 Feb 27];39(11):2010–7. Available from: http://www.healthaffairs.org/doi/10.1377/hlthaff.2020.00980

