## Supplementary figures and images for "Prospective predictive performance comparison between Clinical Gestalt and validated COVID-19 mortality scores"

### Supplementary Figure 1

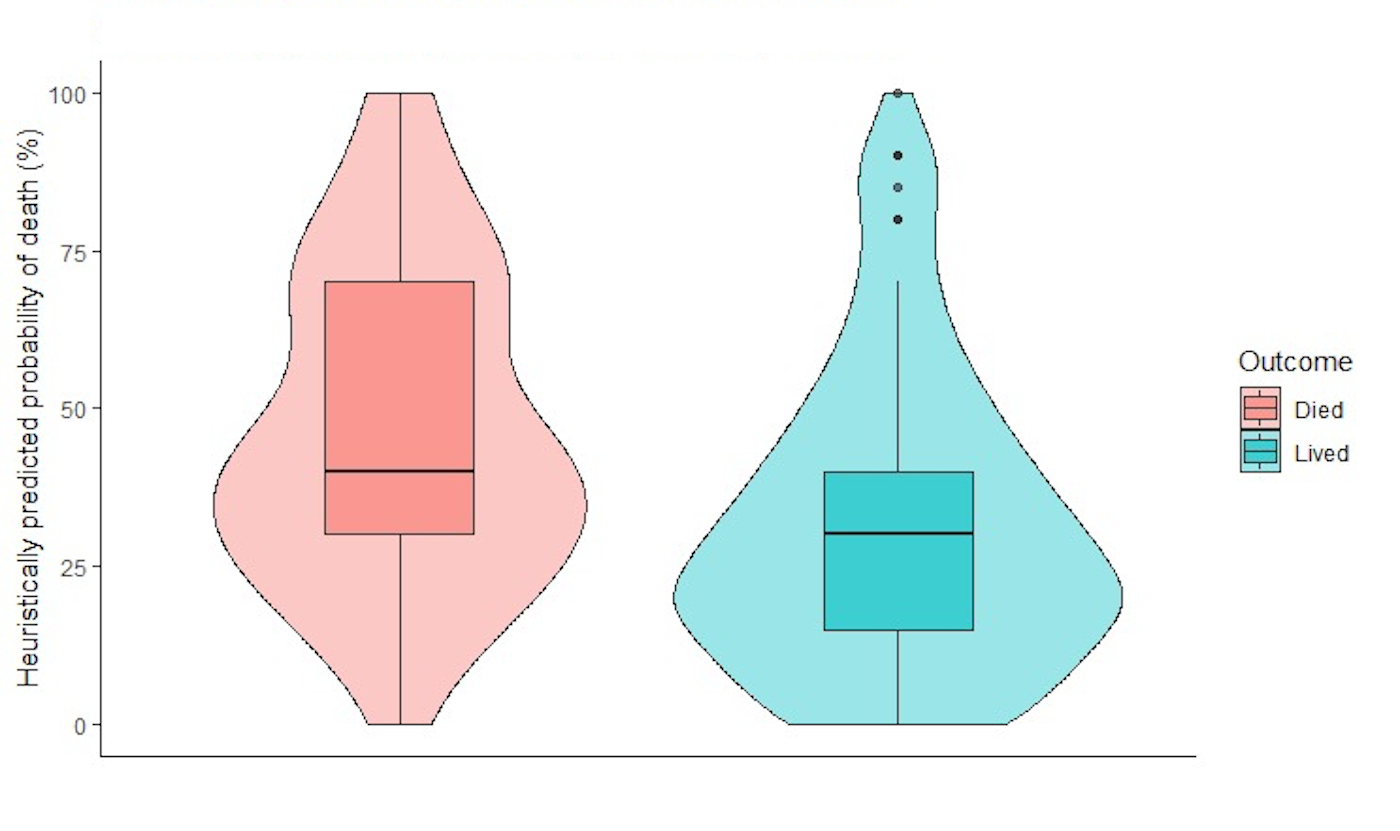
